# Effectiveness of “Reducing Disability in Alzheimer’s Disease” among dyads with moderate dementia

**DOI:** 10.1101/2020.04.07.20056705

**Authors:** Jaime Perales-Puchalt, Kelli Barton, Lauren T Ptomey, C. Michelle Niedens, Amy Yeager, Laura Gilman, Pam Seymour, Amanda George, Susan C. Sprague, Antonio Mirás Neira, Rik Van Dyke, Linda Teri, Eric D Vidoni

## Abstract

Interventions such as Reducing Disability in Alzheimer’s Disease (RDAD) improve the health of care receiver-caregiver dyads but plans to implement it locally in regional community agencies yielded three changes: 1) reduced reliance on licensed clinicians, 2) centralized exercise interventionists and 3) more flexible delivery. We aimed to assess the effectiveness of the Kansas City RDAD implementation (RDAD-KC) among a non-probabilistic sample of dyads with moderate dementia, which addressed these changes. We hypothesized that dyads’ health would improve from baseline to the end-of-treatment. Outcomes improved (p<0.01) from pre to post-intervention: Behavioral symptom severity (range 0-36) decreased from 11.3 to 8.6, physical activity increased from 125.0 to 190.0 minutes/week, caregiver unmet needs (range 0-34) decreased from 10.6 to 5.6, caregiver behavioral symptom distress (0-60) decreased from 15.5 to 10.4 and caregiver strain (0-26) decreased from 11.1 to 9.7. This adapted implementation of RDAD leads to clinically meaningful improvements and might inform scaling-up.

## Introduction

It is well known that the US population is getting older. By 2034, there will be more adults over 65 than children (Vespa, 2018). Older adults and aging policies have emphasized “aging in place” in their own homes and communities (Altshuler & Schimmel, 2010; Oberlink). With this shift in living situations, the US’s current support infrastructure will face unprecedented challenges providing the in-home services and infrastructure that the aging population demands. These challenges especially apply to the growing population of people with dementia (PWD) (Alzheimer’s). Dementia presents additional barriers to providing in-home services due to the increased need for behavioral management and functional support for activities of daily living such as cooking, dressing and hygiene (Riley, Burgener, & Buckwalter).

As outlined in the National Plan to Address Alzheimer’s “enabling family caregivers to continue to provide care while maintaining their own health and well-being” is a primary need for communities and the nation (Strategy 3B(*National Plan to Address Alzheimer’s Disease: 2015 Update*)^66^) (U.S. Department of Health & Human Services, 2015). Achieving this support requires identifying unmet needs and matching these needs with best practices and evidence-based interventions thorough a broad dementia-capable system. Specifically, the plan calls for caregivers to be “linked to interventions shown to decrease burden and depression among caregivers and enhance the care received by people with dementia.” Behavioral challenges and the caregivers’ own health are two of the primary reasons for nursing home placement (Buhr, Kuchibhatla, & Clipp). It has been well established that behavioral challenges are most often associated with care recipient and caregiver dyad factors such as lack of activity, depression, anxiety, and uninformed caregiver approach (Okura et al., 2010; Ornstein & Gaugler, 2012). Compounding the problem, the caregiver/recipient dyad tends to become increasingly isolated through the course of the illness (Buhr et al.; Moyle, Kellett, Ballantyne, & Gracia, 2011). Fatigue, depression, and an unwillingness to expose deficits to a wider community separate them from vital social support (Moyle et al., 2011).

Numerous interventions to improve quality of life have been tested in the home setting for dyads (Dawson, Bowes, Kelly, Velzke, & Ward). Among the most successful has been the Reducing Disability in Alzheimer’s disease (RDAD) (Teri et al., 1998). RDAD is a dyad home-based multicomponent intervention that focuses on exercise for PWD and coping with behavioral symptoms for caregivers. The original goal of RDAD was to decrease behavioral and physical outcomes among PWD and delay their institutionalization. RDAD includes 12 modules on behavior management training and exercise instruction. Behavior management instruction focuses on the “ABCs” of problem behaviors, Antecedents, Behaviors, Consequences, as well as communication skills, pleasurable events and realistic expectations. Exercises include strength, balance, endurance and flexibility. The original dose was comprised of one-hour sessions over three months weekly or biweekly depending on the stage of the intervention. Caregivers are instructed to keep a log with the exercise sessions completed without the instructor and other information to develop coping strategies. The RDAD program initially demonstrated benefits among PWD for depression and increased physical activity and function. Subsequent replication at the state-wide level yielded evidence of the scalability of the program in a in a less research-controlled, more diverse, community environment and effectiveness in reducing caregiver strain and unmet needs (Menne et al., 2014).

According to the National Institutes of Health Intervention Development Model, the intervention development process is incomplete until it is optimally efficacious and implementable with fidelity by practitioners in the community (Onken, Carroll, Shoham, Cuthbert, & Riddle, 2014). In 2016, leaders in the Kansas City aging and disability community (“the Collaborative”) met to discuss potential collaborations. Leadership from a number of organizations participated as group in completing the Community Based Organizations Dementia Capable Quality Assurance Assessment Tool (National Alzheimer’s and Dementia Resource Center, 2020). The survey identified that while most community partners reported providing specialized services for PWD and their caregivers, there was an additional desire for evidence-informed care programs that could be deployed across organizations to leverage training efficiency. The group identified the RDAD as a potential intervention. However, despite the existence of a previous community translation of RDAD, the group identified three major changes to implement RDAD in Kansas City. Changes included 1) not having to rely solely on licensed clinicians given the agency’s shortages, 2) need for greater flexibility in delivery of the content to increase efficiency, and 3) not having to depend on interventionists to deliver exercise education since previous work shows they do not feel comfortable doing so (Menne et al.). We aimed to assess the effectiveness of the Kansas City RDAD implementation (RDAD-KC), which addressed these changes. We hypothesized that dyads’ mental health and physical activity outcomes would improve from baseline to end-of-treatment. We also aimed to assess potential mechanisms by which RDAD-KC might have improved behavioral outcomes among dyads, including physical activity change, length of the intervention and number of completed sessions and modules.

## Methods

### Study design and sample

This pre-post intervention assessment is a secondary analysis of an in-home dementia support services quality improvement project amongst a collaborative of nine community agencies. Agencies administered RDAD-KC to a non-probabilistic sample of dyads they recruited in their service areas. De-identified data was obtained from the partner agencies who agreed to enroll dyads whose eligibility criteria included: living in the community, having moderate dementia (stage 5-6 on the Global Deterioration Scale) (Reisberg, Ferris, de Leon, & Crook, 1982), not having activity restrictions by a physician, being able to walk across a room in their home with or without an assistive device and having a 2.4m^2^ space free of obstacles in which to exercise. Caregivers who did not meet the physical requirements could recruit an “Exercise Buddy” to assist with that component of the program.

### Data source

The Collaborative created a shared data collection workbook that included the necessary pre-and post-intervention outcome measures, basic demographics, compliance work forms from the original RDAD, and agency-modifiable consent, physician notification, and technology agreement. Each agency was encouraged to adapt these documents to their own needs and agency policies. Once client dyads completed their participation in the RDAD-KC program de-identified data was transmitted to the designated Project Evaluator. The project was determined to be secondary data analysis by the Institutional Review Board of the Project Evaluator site, because no identifiable information linked to the outcomes was transmitted outside of the agencies that delivered the intervention, and the outcomes were collected as part of service provided to the community under the cooperative funding agreement.

### Intervention translation

Changes needed to implement RDAD in Kansas City were addressed in the following way. First, to address the need for greater flexibility in delivery of the content, the Collaborative elected to deploy the content as modules that could be delivered to the PWD and caregiver in an order that best fit the dyadic situation, needs, and time available. For example, all 12 lessons could be delivered in 2-12 different sessions. Second, to address the need not to depend on the interventionist to deliver the exercise education, an academic team with exercise delivery experience was recruited to create pre-recorded exercise education videos specific to the original RDAD program that could be viewed via mobile devices (Kindle Fire?) provided to each dyad during the intervention. Four pre-recorded instructional videos that covered the first four behavioral modules (introduction to the program, strengthening exercises, balance exercises, and endurance and flexibility) were created for dyads to view and repeat on their own. Dyads were encouraged to call the exercise interventionist each week during the first four weeks of the intervention to received support, problem solve any barriers to the exercises or to obtaining physical activity, and review possible exercise modifications. The team was also available for phone consultation throughout the remainder of the dyads time in the program, up to 12 weeks. The team was composed of individuals with a Bachelor’s in Exercise Science or equivalent, a clinical license in Occupational Therapy, or an American College of Sports Medicine certification related to exercise prescription or adaptive physical activity. Third, to address the need not to depend on a licensed clinicians to deliver the intervention, community health workers and other experienced individuals would be trained to deliver the intervention as long as they had a bachelor’s degree, at least 100 hours in geriatric experience and dementia training via the Alzheimer’s Association, HRSA Dementia Training Curriculum, or NTG Dementia Capable Care Training.

Consistent with prior implementations was the content. The original RDAD manual, including interventionist materials and caregiver handouts was employed. To incorporate current Alzheimer’s care and research the Alzheimer’s Association publication, Basics of Dementia Care, was substituted for the original RDAD education workbook. Each session of the 12 modules included both behavior management training and exercise instruction. Behavior management instruction focused on the “ABCs” of problem behaviors, Antecedents, Behaviors, Consequences, as well as pleasurable events. Each session of the 12-week program also included exercise training and the dyad was encouraged towards a goal of independently engaging in 30 minutes of exercise most days of the week.

### Outcome Measures

The Collaborative collectively worked with the funding agency to identify outcomes that were specific to caregivers but broadly implementable amongst the various workflows of the partners. Six caregiver-administered outcomes (three caregiver and three PWD-related) were identified as consensus measures that were already being or could be implemented by the partner agencies.

### Caregiver-related outcomes

#### Unmet needs (Measure of Unmet Needs (Gaugler et al., 2004))

The Measure of Unmet Needs is a validated 24-item survey for caregivers that requires “Yes/No” answer regarding additional assistance with categories such as activities of daily living, dementia symptoms, and social support. Outcomes include a summary score of the 24 items and summary scores of each subscales (ADL help, IADL help, dementia symptoms, Timing of care, Formal support, Information and Confidante).

#### Caregiver strain (Modified Caregiver Strain Index (Thornton & Travis, 2003))

The Modified Caregiver Strain Index is a validated 13-item screener for caregiver strain after hospital discharge of an older adult family member. For each of the items, the caregiver can respond either no (0), yes sometimes (1), or yes on a regular basis (2). Outcomes include an overall summary score as well as four factors informed by a previous factor analysis: Inconvenience, Adjustment, Upsetting and Work adjustments (Rubio, Berg-Weger, & Tebb, 1999).

#### Behavioral symptom-related distress (Neuropsychiatric Inventory Questionnaire (NPI-Q) (Kaufer et al., 2000))

The Neuropsychiatric Inventory (NPI) is a validated clinical instrument for evaluating psychopathology in dementia. If any of the 12 neuropsychiatric symptoms is present, caregivers rate their own distress on a six-point scale. An overall distress summary score is calculated by adding the distress scores of all items.

### PWD-related outcomes

#### Behavioral symptom-related severity (Neuropsychiatric Inventory Questionnaire (NPI-Q) (Kaufer et al., 2000))

If any of the 12 neuropsychiatric symptoms is present, caregivers rate the level of severity for the PWD on a three-point scale. An overall severity summary score is calculated by adding the severity scores of all items. Outcomes also include summary scores of two factors informed by a previous factor analysis: Negative/oppositional and Anxiety/restlessness (Travis Seidl & Massman, 2016).

#### Sedentary behavior (Physical Activity (Teri et al.))

Interventionists asked about the number of days a PWD spent either sitting or lying down in the last week. These ratings were subjective caregiver responses.

#### Physical activity (Physical Activity (Teri et al.))

Interventionists asked about the number of minutes the PWD was physically active (including walking and cycling but excluding wandering or pacing) during the last week.

### Statistical Analyses

The final analytical sample included those eligible for the intervention, who had a family caregiver, had no intellectual disability and completed nine out of 12 modules (>75%) of the intervention. Descriptive statistics were calculated for baseline characteristics.

Effectiveness analysis: Since the distribution of overall scores of all caregiver outcomes, behavioral symptom distress subscales and behavioral symptom severity of the PWD was normal, we used paired-samples t-tests to assess their pre-post-intervention change. The distribution of unmet needs subscales and caregiver strain subscales as well as physical activity and sedentary behavior was not normal. Therefore, we analyzed the change in these variables using paired samples Wilcoxon for all variables except for the “Work adjustments” subscale of the Caregiver Strain Index, for which we used a McNemar test due to its binary distribution. Mechanistic analysis: Given the normal distribution of the change in all overall outcome scores, we calculated Pearson correlations to analyze between-outcome associations. The number of completed RDAD sessions, modules and length of the intervention in days were all positively skewed. For this reason, we analyzed their association with change in overall outcome scores using Spearman correlations. We used SPSS v20.0 for all data analyses and a significance of p<.05.

## Results

Among the 157 screened dyads, 47 were excluded for this secondary analysis due to intellectual or developmental disability (an alternative diagnosis for the program). The analysis also excluded three dyads that included a formal caregiver and another three in which the relationship between the caregiver and PWD was not defined. Among the remaining 104 dyads, 38 did not complete at least nine RDAD modules and were excluded from the analysis (Table 1). PWD who completed at least nine modules were more likely to be women than non-completers (66.6% vs 36.8%, p<0.05) and were less likely to live alone (7.0% vs 24.3%, p<0.05). Caregivers who completed at least nine modules were less likely to be women than non-completers (58.5% vs 78.4%, p<0.05).

**Table 1.** Participant characteristics at baseline.

|  | Completers |  |  |  | Non-completers |  |  |  |
| --- | --- | --- | --- | --- | --- | --- | --- | --- |
| Characteristics | Persons with dementia with informal caregiver; n=66 |  | Informal caregivers; n=66 |  | Persons with dementia with informal caregiver; n=38 |  | Informal caregivers; n=38 |  |
|  | % or M | SD or N | % or M | SD or N | % or M | SD or N | % or M | SD or N |
| Age in years | 77.4 | 9.4 | 68.5 | 12.9 | 77.9 | 11.2 | 69.9 | 8.8 |
| Women | **66.6** | **40** | **58.5** | **38** | **36.8** | **14** | **78.4** | **29** |
| Served in military | 16.7 | 11 | 17.2 | 11 | 28.6 | 10 | 5.6 | 2 |
| Rural setting | 17.0 | 9 | 14.3 | 7 | 31.6 | 12 | 27.3 | 6 |
| In-home wireless capability | 85.5 | 53 | - | - | 74.3 | 26 | - | - |
| Ethnoracial group |  |  |  |  |  |  |  |  |
| White | 72.9 | 35 | 77.1 | 37 | 78.1 | 25 | 80.6 | 25 |
| African American | 20.8 | 10 | 20.8 | 10 | 15.6 | 5 | 16.1 | 5 |
| Latino | 4.2 | 2 | 2.1 | 1 | 6.3 | 2 | 3.2 | 1 |
| Asian | 2.1 | 1 | 0.0 | 0 | - | - |  |  |
| Relationship to caregiver |  |  |  |  |  |  |  |  |
| Partner | 71.2 | 47 | - | - | 68.4 | 26 | - | - |
| Child | 19.7 | 13 | - | - | 23.7 | 9 | - | - |
| Other informal | 9.1 | 6 | - | - | 7.9 | 3 | - | - |
| Lives alone | **7.0** | **4** | - | - | **24.3** | **9** | - | - |

Rural: adjacent to population < 50,000; bold: completers vs non-completers p value<0.05 for t-test or $\chi^2$

The final analytic sample included 66 dyads. PWD’s mean age was 77.4 years and ranged from 52 to 95. Among the PWD, 16.7% were veterans, 17.0% lived in rural areas and 85.5% had in-home Wi-Fi. PWD mostly identified as non-Latino White (72.9%) or non-Latino African American (19.7%), and their caregiver was usually their partner (71.2%) or their child (19.7%). Caregivers’ mean age was 68.5 years and ranged from 33 to 90. Among the caregivers, 17.2% were veterans and 14.7% lived in rural areas.

Caregivers mostly identified as non-Latino White (77.1%) or non-Latino African American (20.8%). Dyads completed a median of six sessions (min 3, max 20), 49.9% completed the 12 modules and the median number of days from the first to the last RDAD session was 70 (min 28, max 219).

Table 2 shows the effectiveness of RDAD-KC on PWD behavioral symptom severity, physical activity, and sedentary behavior. The overall mean of PWD behavioral symptom severity, as rated by the caregiver in the NPI-Q, decreased from 11.3 to 8.6 (p<0.001) as did the subscale symptoms of negative/oppositional behavior (6.0 to 4.7, p<0.01) and anxiety/restlessness (4.1 to 3.0, p<0.001). The median minutes per week physical activity increased from 125.0 to 190.0 (p<0.01) but the days per week spent sitting or lying did not change significantly.

**Table 2.** Effectiveness: change in means from baseline to follow up for person with dementia outcomes among completers.

|  | Persons with dementia |  |  |  |  |  |  |
| --- | --- | --- | --- | --- | --- | --- | --- |
|  | N | Baseline |  | Follow up |  |  |  |
| **Person with dementia** |  | M/Q2 | SD/Q3<br>-Q1 | M/Q2 | SD/Q3<br>-Q1 | t/z | P-Value |
| NPI-Q Severity (0-36) <sup>a</sup> | 58 | 11.3 | 7.1 | 8.6 | 6.9 | 3.8 | **0.000** |
| Negative/oppositional <sup>a</sup> | 60 | 6.0 | 4.1 | 4.7 | 3.9 | 3.2 | **0.003** |
| Anxiety/restlessness <sup>a</sup> | 63 | 4.1 | 2.7 | 3.0 | 2.6 | 3.9 | **0.000** |
| Physical activity<br>(min/wk) <sup>b</sup> | 62 | 125.<br>0 | 222.5 | 190.<br>0 | 290.0 | -3.0 | **0.003** |
| Sat or laid most of the<br>day (days/wk) <sup>b</sup> | 62 | 0.0 | 1.0 | 0.0 | 1.0 | -0.2 | 0.824 |

NPI-Q: Neuropsychiatric Inventory Questionnaire, Paired samples t-test <sup>a</sup> and Wilcoxon <sup>b</sup>

Table 3 shows the effectiveness of RDAD-KC on caregiver unmet needs, behavioral symptoms-related distress and strain. The mean number of unmet needs decreased from 10.6 at baseline to 5.6 at follow up (p<0.001). The median number of all unmet need subscales also decreased statistically from baseline to follow up (p<0.05). The overall mean of caregiver distress associated with behavioral symptoms, as rated in the NPI-Q, decreased from 15.5 to 10.4 (p<0.001) as did the overall caregiver strain, which decreased from 11.1 to 9.7 (p<0.010). The “upsetting” subscale was the only caregiver strain subscale that decreased statistically (p<0.010). No correlation between change in physical activity of the PWD and overall outcomes (behavioral symptom severity and distress, unmet needs or caregiver strain) was statistically significant. Change in unmet needs was the only outcome associated with change in sedentary behavior (0.469).

**Table 3.** Effectiveness: change in means from baseline to follow up for caregiver outcomes among completers.

|  | Informal caregivers |  |  |  |  |  |  |
| --- | --- | --- | --- | --- | --- | --- | --- |
|  | N | Baseline |  | Follow up |  |  |  |
| Caregiver |  | M/Q2 | SD/Q3-Q1 | M/Q2 | SD/Q3-Q1 | T/z | P-Value |
| Unmet needs (0-34) <sup>a</sup> | 62 | 10.6 | 6.2 | 5.6 | 6.2 | 6.0 | **0.000** |
| ADL <sup>b</sup> | 63 | 0.0 | 1.0 | 0.0 | 0.0 | -3.2 | **0.001** |
| IADL <sup>b</sup> | 57 | 1.0 | 2.0 | 0.0 | 1.0 | -2.5 | **0.011** |
| Dementia symptoms <sup>b</sup> | 65 | 1.0 | 1.0 | 0.0 | 1.0 | -4.9 | **0.000** |
| Timing of care <sup>b</sup> | 63 | 1.0 | 2.0 | 0.0 | 1.0 | -2.1 | **0.037** |
| Formal support <sup>b</sup> | 59 | 3.0 | 4.0 | 1.0 | 2.3 | -4.5 | **0.000** |
| Information <sup>b</sup> | 61 | 1.0 | 2.0 | 0.0 | 1.0 | -3.9 | **0.000** |
| Confidante <sup>b</sup> | 61 | 1.0 | 2.8 | 0.0 | 1.0 | -3.2 | **0.002** |
| NPI-Q Distress (0-60) <sup>a</sup> | 58 | 15.5 | 9.9 | 10.4 | 9.0 | 5.2 | **0.000** |
| Caregiver Strain Index (0-26) <sup>a</sup> | 64 | 11.1 | 5.0 | 9.7 | 5.3 | 2.9 | **0.006** |
| Inconvenience <sup>b</sup> | 63 | 4.0 | 2.0 | 4.0 | 3.0 | -1.2 | 0.229 |
| Adjustment <sup>b</sup> | 63 | 2.0 | 2.0 | 2.0 | 1.0 | -1.7 | 0.092 |
| Upsetting <sup>b</sup> | 64 | 2.0 | 0.0 | 2.0 | 1.0 | -3.3 | **0.001** |
| Work adjustments; %<br>n <sup>c</sup> | 65 | 36.8 | 25 | 31.3 | 21 | - | 0.388 |

NPI-Q: Neuropsychiatric Inventory Questionnaire, Paired samples t-test <sup>a</sup>, Wilcoxon <sup>b</sup> and McNemar <sup>c</sup>

**Table 4.** Mechanistic analysis: between-outcome correlations and correlations between intervention implementation outcomes and effectiveness outcomes.

|  | Unmet needs change | NPI-Q Distress change | Caregiver Strain Index change | NPI-Q Severity change | Physical activity change | Sat or laid most of the day change | Number of sessions | Number of modules | Days from beginning to end of treatment |
| --- | --- | --- | --- | --- | --- | --- | --- | --- | --- |
| Unmet needs change | 1 | - | - | - | - | - | - | - | - |
| NPI-Q Distress change | 0.217 | 1 | - | - | - | - | - | - | - |
| Caregiver Strain Index change | **0.450** | **0.425** | 1 | - | - | - | - | - | - |
| NPI-Q Severity change | 0.101 | **0.851** | **0.438** | 1 | - | - | - | - | - |
| Physical activity change | 0.001 | 0.020 | 0.005 | -0.100 | 1 | - | - | - | - |
| Sat or laid most of the day change | **0.468** | 0.005 | 0.066 | 0.021 | -0.121 | 1 | - | - | - |
| Days from beginning to end of treatment | -0.094 | 0.047 | 0.214 | -0.022 | 0.141 | -0.121 | 1 | - | - |
| Number of sessions | **-0.325** | -0.091 | 0.014 | -0.124 | -0.045 | -0.112 | **0.634** | 1 | - |
| Number of modules | 0.167 | -0.045 | 0.134 | -0.029 | 0.148 | -0.090 | **0.490** | 0.242 | 1 |

NPI-Q: Neuropsychiatric Inventory Questionnaire; **Bold:** p<0.05

Change in caregiver strain was statistically associated with change in caregiver unmet needs (0.450) and in distress related to behavioral symptoms (0.425) as well as in severity of behavioral symptoms (0.438). Change in caregiver distress related to behavioral symptoms was also associated with change in severity of behavioral symptoms (0.851). The number of days from the beginning to the end-of-treatment was associated with the number of sessions (0.634) and number of modules completed (0.490). The only outcome associated with any of these implementation variables was the number of RDAD sessions, which associated with the change in unmet needs (−0.325).

## Discussion

Caregiver support and dementia capable care amongst America’s service providing agencies is a critical need (U.S. Department of Health & Human Services). In an attempt to address this need in Kansas City, the Collaborative selected and modified the proven RDAD intervention for semi-standardized training and implementation across the agencies. The agencies found that the RDAD-KC improved caregiver unmet needs, behavioral symptom distress and strain as well as the PWD’s behavioral symptom severity and their level of physical activity. It is important to note that these improvements are taking place in the moderate stage of dementia, a time when behavioral symptoms and challenges faced by caregivers, usually begin to escalate. Additionally, this implementation showed that the more DAD-KC sessions that were attended, the more caregiver unmet needs were addressed.

To our knowledge meaningful clinical differences in the Measure of Unmet Needs and the Modified Caregiver Strain Index have not been established. However, our analyses showed a 47% decrease in the number of unmet caregiver needs, suggesting that the RDAD-KC program, interaction with supports in the home and tele-exercise counseling improved access to resources. Similarly, caregivers reported a 12.6% reduction in feelings of strain. Differences in Neuropsychiatric symptom severity and distress scales suggest that in-home support and the RDAD-KC meaningfully reduced behavioral symptoms. NPI-Q severity was reduced by 2.7, close to the 2.77 points reported to be a minimal clinically significant difference (Mao, Kuo, Huang, Cummings, & Hwang). The reduction of the NPI-Q distress score, related to the amount of distress the caregiver experiences, was reduced by 5.1 points, exceeding the reported minimal clinically significant difference of 3.1 (Mao et al.).

Caution is warranted when considering these results, as limitations of this study include a pre-post design with no control group, which prevents from causation inference. The outcomes were not collected as part of a prospective research study. Rather, the outcomes and data collection were part of a mutually agreed upon standard set in a collaborative service provision project in Kansas City. Significant flexibility was afforded to the agencies in the delivery of the intervention and the integration of the outcomes into workflows. Thus, these results reflect “real world” realities, essential to our goals.

The results do provide support that several otherwise independent agencies can mutually deliver a semi-standardized dementia support program in the home and identify meaningful improvements for the caregiver and PWD. The number of dyads who could not complete the intervention is problematic, though perhaps not unexpected. The requirements of the project and the cooperative funding agreement necessitated the inclusion of PWD who were at moderate stage dementia. Moderate stage dementia is typically a transitional time, when caregivers begin to consider placement. Unfortunately, the service providing agencies did not collect information on reasons for ending participation in the intervention. Post hoc interviews with the agencies suggest that caregiver burden, medical crisis, and residential care placement were the primary reasons for ending program participation. This was a Quality Improvement project and research was not its primary goal. For this reason, results are specific to the partnering organizations and are not necessarily generalizable to other communities or collaborative groups.

This study has several clinical implications. This study highlights the need to tailor interventions to the community. RDAD-KC, a less stringent version of RDAD led to dyad clinically significant benefits that are comparable to original RDAD results. The association between number of RDAD-KC sessions and unmet needs suggests that while a degree of flexibility in the implementation of RDAD-KC is important, interventionists should encourage dyads to complete as many sessions as feasible. RDAD-KC participants did not improve their levels of sedentary behavior. Further implementations might need to stress the need to reduce sedentary behavior given its link with mental and physical health (de Rezende, Lopes, Rey-Lopez, Matsudo, & do Carmo Luiz, 2014).

## Conclusion

This implementation of RDAD leads to clinically meaningful improvement of behavioral symptom distress among caregivers and severity among PWD. RDAD-KC also leads to improvement in other outcomes including caregiver strain and unmet needs and PWD physical activity. Findings from this study provide useful real-life implementation information to address the need to enable family caregivers to continue to provide care while maintaining their own health and well-being identified in the National Plan to Address Alzheimer’s. Despite the uniqueness of every community, the RDAD-KC implementation experience may contribute to informing the scaling-up of RDAD in other communities.

## Data Availability

The data that support the findings of this study are available from the corresponding author, upon reasonable request.

## Acknowledgements

The corresponding author affirms that he has listed everyone who contributed significantly to the work and has obtained written consent from all contributors who are not authors and are named in the Acknowledgment section. The authors thank all community agencies who participated in the RDAD-KC project, the ACL officers for their input during the project and all individuals who participated in RDAD-KC.

